# Predicting infectious disease for biopreparedness and response: A systematic review of machine learning and deep learning approaches

**DOI:** 10.1101/2022.06.30.22277117

**Authors:** Ravikiran Keshavamurthy, Samuel Dixon, Karl T. Pazdernik, Lauren E. Charles

**Affiliations:** Pacific Northwest National Laboratory, Richland, WA 99354, USA; Paul G. Allen School for Global Health, Washington State University, Pullman, WA 99164, USA; Department of Statistics, North Carolina State University, Raleigh, NC 27695, USA

## Abstract

Despite the complex and unpredictable nature of pathogen occurrence, substantial efforts have been made to better predict infectious diseases (IDs). Following PRISMA guidelines, we conducted a systematic review to investigate the advances in ID prediction capabilities for human and animal diseases, focusing on Machine Learning (ML) and Deep Learning (DL) techniques. Between January 2001 and May 2021, the number of relevant articles published steadily increased with a significantly influx after January 2019. Among the 237 articles included, a variety of IDs and locations were modeled, with the most common being COVID-19 (37.1%) followed by Influenza/influenza-like illnesses (8.9%) and Eastern Asia (32.5%) followed by North America (17.7%), respectively. Tree-based ML models (38.4%) and feed-forward DL neural networks (26.6%) were the most frequent approaches taking advantage of a wide variety of input features. Most articles contained models predicting temporal incidence (66.7%) followed by disease risk (38.0%) and spatial movement (31.2%). Less than 10% of studies addressed the concepts of uncertainty quantification, computational efficiency, and missing data, which are essential to operational use and deployment. Our study summarizes the broad aspects and current status of ID prediction capabilities and provides guidelines for future works to better support biopreparedness and response.

## Introduction

Infectious disease (ID) events have plagued human and animal populations throughout history, resulting in massive numbers of morbidities and mortalities as well as substantial social and economic impacts across the world^1^. The effects of climate change, urbanization, and globalization have rendered these diseases borderless, enabling them to spread easily across regions and inevitably increasing the risk of epidemics and pandemics. Currently, ID prediction is one of the most important operational epidemiological tools with the potential to provide early warning to actively prevent disease occurrence and spread. By combining robust data collection, engineering, and analysis strategies, predicting disease event information, such as location, timing, intensity, and various other factors responsible for its occurrence, becomes possible. Timeliness of this type of predicted information is crucial for decision makers to effectively mobilize health resources to the area of concern and properly implement control and prevention strategies^2^.

Predicting ID is a challenging task mainly due to the complex and unpredictable nature of pathogen ecology and evolutionary dynamics^3^. These inherent complexities demand robust uncertainties quantification systems for better decision making. The challenge of ID prediction is exacerbated due to inadequate and biased disease surveillance initiatives, a lack of disease reporting systems, as well as incomplete and delayed epidemiological data sharing^4,5^. Despite these limitations, significant efforts have been made in the past couple of decades to utilize ID prediction models in operational control and prevention strategies. In particular, the emergence of the coronavirus disease 2019 (COVID-19) pandemic has resulted in accelerated development and integration of ID prediction models in worldwide public health decision making^6^.

In recent years, factors, such as an exponential increase in computing power, easy access to large and diverse datasets, and advancements in artificial intelligence, have facilitated extraordinary growth in the field of infectious disease predictions^7^. Machine Learning (ML) and Deep Learning (DL) methods are widely used for a variety of disease intelligence tasks, including temporal, spatial, and risk factor predictions^8^. ML models have been shown to outperform traditional statistical techniques to give more accurate and reliable predictions^9,10^. The popular ML techniques most widely used in the field of ID prediction include tree-based approaches ^10–12^ and Support Vector Machines (SVM)^13–15^ due to their ease of implementation and interpretability. On the other hand, DL techniques, such as feed-forward neural networks (FNN)^16,17^ and recurrent neural networks (RNN)^18,19^, are popular for their ability to integrate large and complex data into their predictions.

There are many, complex factors that contribute to and influence the presence of an ID event, such as epidemiologic, geographic, climatic, demographic, behavioral, and sociopolitical. Traditional ID prediction models can only process a limited number of explanatory variables and do not perform will on cross-correlated features. On the other hand, ML and DL models excel at processing large amounts of feature data and finding complex and hidden connections amongst data sources. ID prediction modeling has, therefore, greatly benefited from the recent “big data” revolution^20^. Remote sensing satellite imagery and census data yield high resolution information about critical disease related factors, such as climate, environment, population density, and demography. With the increase in worldwide internet and mobile phone usage, non-traditional information (e.g., internet searches, social media usage, phone call records, news media trends, and population mobility data) are also readily available. The ML and DL approaches have become highly efficient in utilizing large and complex information gathered through multiple channels to provide a unique opportunity to understand and model ID dynamics like never before^3,21^. However, the utilization of large datasets and increased complexity of the prediction models could lead to an exponential rise in computational requirements. Hence, optimizing the memory and processing requirements of the ML and DL algorithms without compromising their predictive capabilities is crucial.

This study investigates the advances to and quality of ID prediction capabilities, focusing on ML and DL techniques applied over the past two decades. To do this evaluation, we systematically reviewed the scientific literature to identify research that included ML and/or DL models to predict IDs in humans and/or animals. Within the collection, we highlighted specific tasks performed by each prediction model type, input features used for model building, the study spatial and temporal scales, and error metrics applied. We specifically noted if the studies addressed the important issues of uncertainty quantification, computational efficiency, and missing data when building the models. By focusing on the above-mentioned research areas, we identified the best approaches and strategies as well as revealed gaps present in the field of ID prediction modeling. This systematic analysis can be used as a guide to improve future research studies, to better address operational needs for model deployment, and to inform areas where public health and veterinary policies can help improve predictive capabilities.

### Methodology

To assess the application of ML and DL techniques in the field of infectious disease prediction, we conducted a systematic review following Preferred Reporting Items for Systematic Reviews and Meta-Analyses (PRISMA) guidelines^22^. A diverse set of subject matter experts, spanning infectious diseases, public health, epidemiology, computer engineering, data science, and statistics, formulated the following specific research questions:

- Which IDs are modeled using ML and DL techniques?
- Which global geographic regions are modeled in the ID prediction studies?
- What is the trend and extent of ML and DL types and sub-types used in ID predictions?
- What are the various tasks performed by ID prediction models?
- What are the different input features used for ID predictions?
- What are the spatial (geographic extent) and temporal (duration) scales of the studies?
- What are the error metrics used?
- Is uncertainty quantification, computational efficiency, or missing data handling addressed?

### Eligibility criteria

Specific eligibility criteria were developed based on subject matter expert recommendations. Inclusion criteria required that the study (1) must explicitly include temporal, spatial, and/or risk prediction models of infectious diseases; (2) must utilize ML and/or DL techniques for predictions; (3) must be an original study; and (4) must be published in a peer-reviewed journal in the English-language between Jan 2001 and May 2021. We excluded prediction studies containing sexually transmitted diseases, cancer, clinical trials, and only biomarker data (e.g., genomics, proteomics, transcriptomics). In addition, we did not include research that primarily utilized traditional statistics-based regression or classification methods (e.g., linear, non-linear, autoregressive moving average, logistic or Poisson models). Preprints, book chapters, conferences presentations, reviews, opinions, commentaries, and dissertations were excluded. We also excluded articles with missing or inaccessible full texts.

### Search strategy

In May 2021, the scientific literature databases of PubMed, Web of Sciences, Embase, Scopus, and Google Scholar were searched to guarantee effective and adequate coverage of targeted studies (Table 1). The literature published between Jan 2001 and May 2021 was searched using the keywords recommended by subject matter experts. We restricted the Google Scholar searches to the first 300 results, which provides an acceptable search coverage of academic literature without excluding useful references^23^. The citation manager Mendeley (https://www.mendeley.com/) was used to manage imported review citations.

**Table 1:**
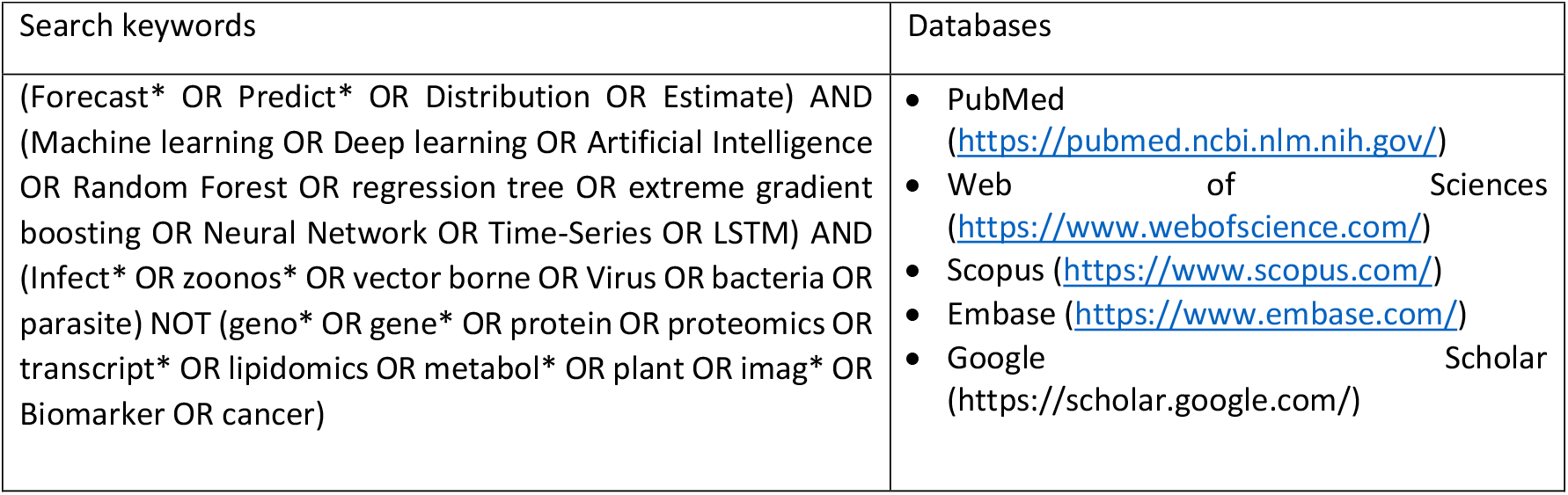
Search keywords and scientific literature databases used to identify potentially relevant publications for systematic review.

### Selection Strategy

Citations were first de-duplicated before proceeding to the manual screening of abstracts. As the first step, each abstract was evaluated by two independent reviewers for possible eligibility in the systematic review based on defined eligibility criteria. Next, the full texts of potential candidate articles were evaluated in detail by the reviewers to ensure all criteria were met. Articles that passed the two-part screening were included in the final publication list and, ultimately, in the systematic review.

### Information extraction

The ML and DL models present in the review literature were classified into broad categories based on the tasks they performed listed below.

### Temporal prediction models

utilize historic disease information to predict future disease events. These models attempt to answer *when the next disease outbreak would occur* in the future based on past events.

### Spatial prediction models

utilize historic disease information to predict the geographic distribution of disease events. These models attempt to answer *where the next disease outbreak might occur* by imputing the locations where disease occurrence information is not available.

### Risk prediction models

assess the relationship between disease events and various factors associated with their occurrence. These models attempt to *estimate spatial and/or temporal risk factors* correlated with the disease event.

During the process of full-text review, the reviewers recorded the following information: model types and subtypes, disease names, primary study hosts, input features or explanatory variables used for predictions, study area, study duration, temporal forecasting distance, error metrics used, uncertainty quantification, missing data handling, and computational efficiency. These groupings are not mutually exclusive. For example, Zhang et al^a211^ compared the performance of temporal prediction models belonging to FFNN and RNN to forecast typhoid fever incidence in China. To evaluate their model performance, they used three error metrics (mean absolute error, mean absolute percentage error, and mean square error). Hence, this citation was placed under multiple prediction model subtype and error metric categories. Similarly, if a publication model performed multiple tasks, such as modeling multiple diseases, geographic locations, or prediction categories, the citation was placed in all relevant categories. Any differences in opinion between the independent reviewers raised during the collection, screening, and information recording processes of the review were resolved through internal discussion until consensus was achieved.

## Results

We identified 16,148 articles that were published in peer reviewed journals between January 2001 and May 2021 (Fig. 1). After removing the duplicates and screening the records for inclusion and exclusion criteria, 237 articles were selected for the final systematic review. The complete list of articles that were included in this systematic review is provided in Supplementary Note (a1–a237).

**Figure 1:**
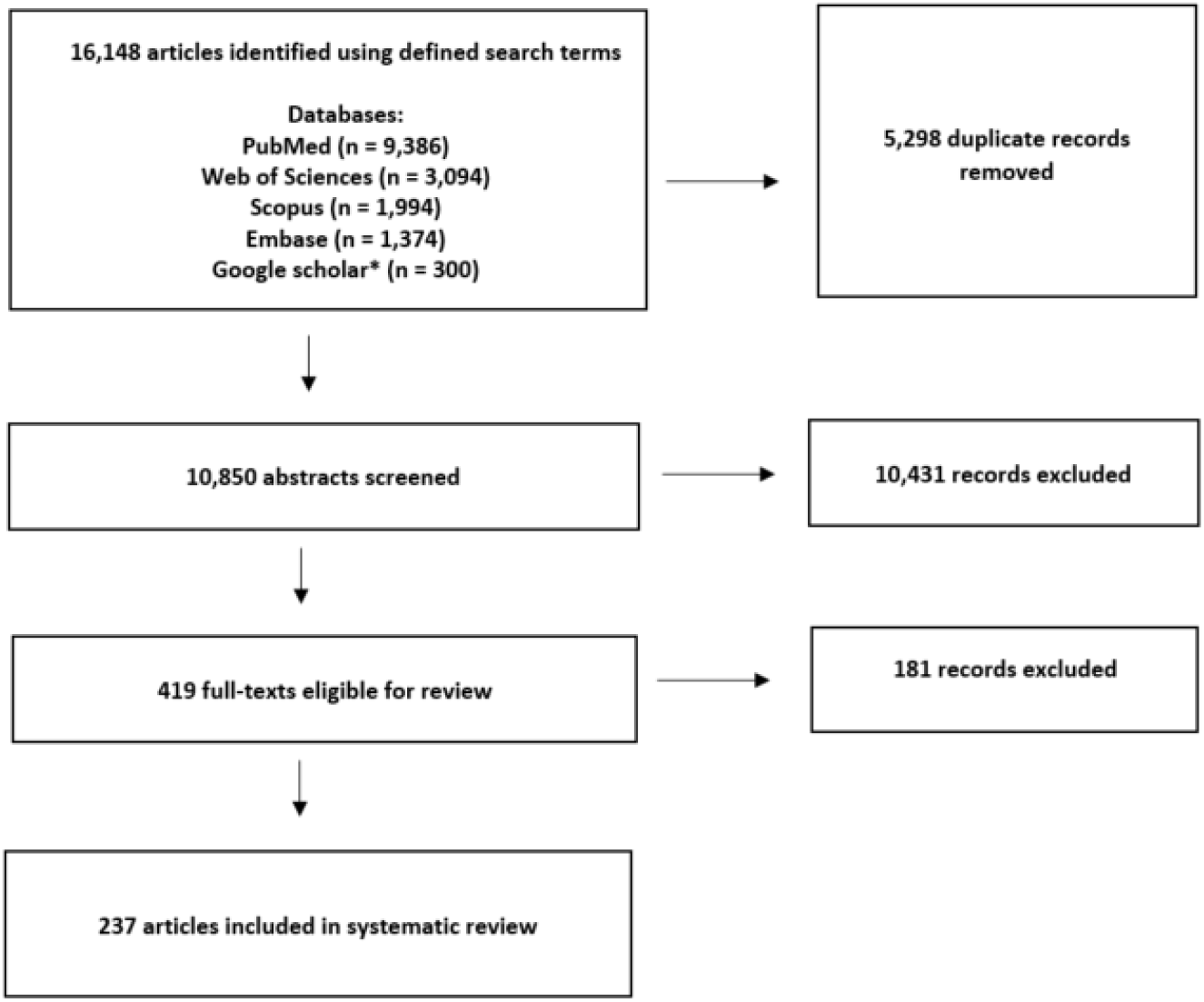
PRISMA flow diagram. The illustration of the overall the selection process. * Google Scholar searches were restricted to the first 300 results

### ML and DL modeling for infectious disease prediction

Among the large and diverse number of ID prediction models applying ML or DL methods found in the literature based on our criteria, COVID-19 undoubtedly received the most attention and was studied in 88 (37.1%) articles. Influenza and influenza-like illnesses were modeled in 22 (9.3%) articles followed by dengue and malaria in 21 (8.9%) and 12 (5.1%) articles, respectively. The complete list of all the infectious diseases identified in the literature review along with their citations is presented in Table 2.

**Table 2:** Citations categorized by infectious disease and study host

| Disease | Study host | No. of articles |
| --- | --- | --- |
| COVID-19 | Human<sup>a1-a88</sup> | 88 |
| Influenza and influenza-like illnesses | Human<sup>a89-a110</sup> | 22 |
| Dengue | Human<sup>a111-a130</sup>, vector<sup>a131</sup> | 21 |
| Malaria | Humana<sup>132-a142</sup>, vector<sup>a143</sup> | 12 |
| Tuberculosis | Human<sup>a138, a144-a153</sup> | 11 |
| Other mosquito-borne diseases* | Human<sup>a113, a155, a156, a158, a159</sup>, wild birds<sup>a154, a157</sup>, horse<sup>a157</sup>, vector<sup>a157</sup> | 8 |
| Avian influenza | Poultry<sup>a160, a165-a167</sup>, human<sup>a162-a164</sup>, wild birds<sup>a160-a161</sup> | 8 |
| Tick-borne diseases | Human<sup>a168-173</sup>, vectors<sup>a172-a175</sup> | 8 |
| Non-specific diseases** | Humans<sup>a182-188</sup>, livestock<sup>a184</sup>, wildlife<sup>a184</sup> | 7 |
| Brucellosis | Human<sup>a176-a181</sup> | 6 |
| Hand foot and mouth disease (HFMD) | Humans<sup>a189-193</sup> | 5 |
| Hepatitis A, B, or E | Humans<sup>a176, a194-a197</sup> | 5 |
| Leishmaniasis | Humans<sup>a200-a202</sup>, dogs<sup>a198</sup>, vectors<sup>a199</sup> | 5 |
| Hemorrhagic fever with renal syndrome (HFRS) | Humans<sup>a176, a203-a205</sup> | 4 |
| Plague | Wild animals<sup>a207-209</sup>, humans<sup>a206</sup>, domestic animals<sup>a209</sup>, vectors<sup>a208</sup> | 4 |
| Typhoid | Humans<sup>a120, a138, a176, a210</sup> | 4 |
| Anthrax | Humans<sup>a211-a213</sup>, livestock<sup>a211-a213</sup>, wild animals<sup>a212-a213</sup> | 3 |
| Zika | Humans<sup>a218-a220</sup> | 3 |
| Ebola and Marburg | Humans<sup>a214-a215</sup>, wild animals<sup>a214</sup> | 2 |
| Hantavirus | Humans<sup>a216-a217</sup> | 2 |
| Others*** | Humans<sup>a105, a135, a138, a176, a221-a231</sup>, domestic animals<sup>a232-236</sup>, wild animals<sup>a237</sup> | 21 |

\*Other mosquito-borne diseases included West Nile fever, lymphatic filariasis, yellow fever, onchocerciasis, chikungunya
\*\*Non-specific diseases included antibiotic resistance, infectious diarrhea, emerging zoonotic diseases, foodborne disease, acute respiratory infectious disease
\*\*\*Others included scarlet fever, chickenpox, bacillary dysentery, cholera, cryptosporidiosis, schistosomiasis, whooping cough, porcine reproductive and respiratory syndrome, salmonella infection, *E. Coli* infection, leptospirosis, porcine epidemic diarrhea, African swine fever, rabies, peste des petits ruminants. Note: if an article included models for multiple diseases and primary study hosts, it was placed in each respective category.

A large majority (205, 86.5%) of articles focused on modeling only humans followed by only domestic animals (9, 3.8%), only wildlife (6, 2.5%), and only vectors (6, 2.5%) (Fig. 2). There were only a few articles (12, 5.1%) that used more than one host species for modeling IDs.

**Figure 2:**
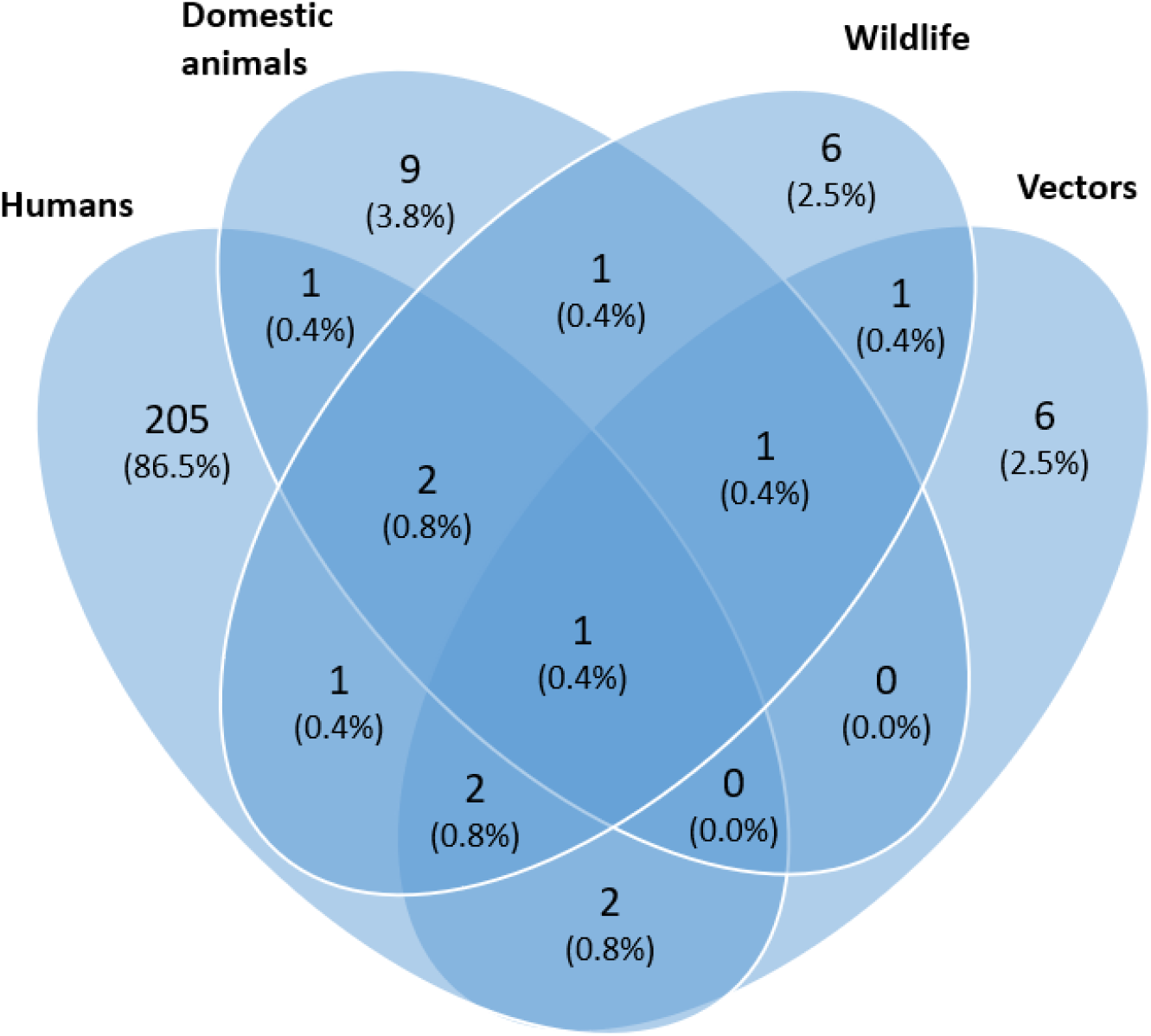
Venn diagram of articles grouped by host species included in infectious disease modeling using machine learning and deep learning techniques. Domesticated animals include livestock and companion animals; wildlife includes wild animals and birds.

### Regional distribution of studies

Of the 237 included studies, the majority of them were focused on Eastern Asia (77, 32.5%), followed by North America (42, 17.7%), Southern Asia (31, 13.1%), Latin America (20, 8.4%) and Western Europe (18, 7.6%). There were 36 (15.2%) studies that included multiple regions (more than four) which were grouped as a separate category. A complete breakdown of the articles with ID models belonging to each geographical region grouped by diseases is presented in Fig. 3.

**Figure 3:**
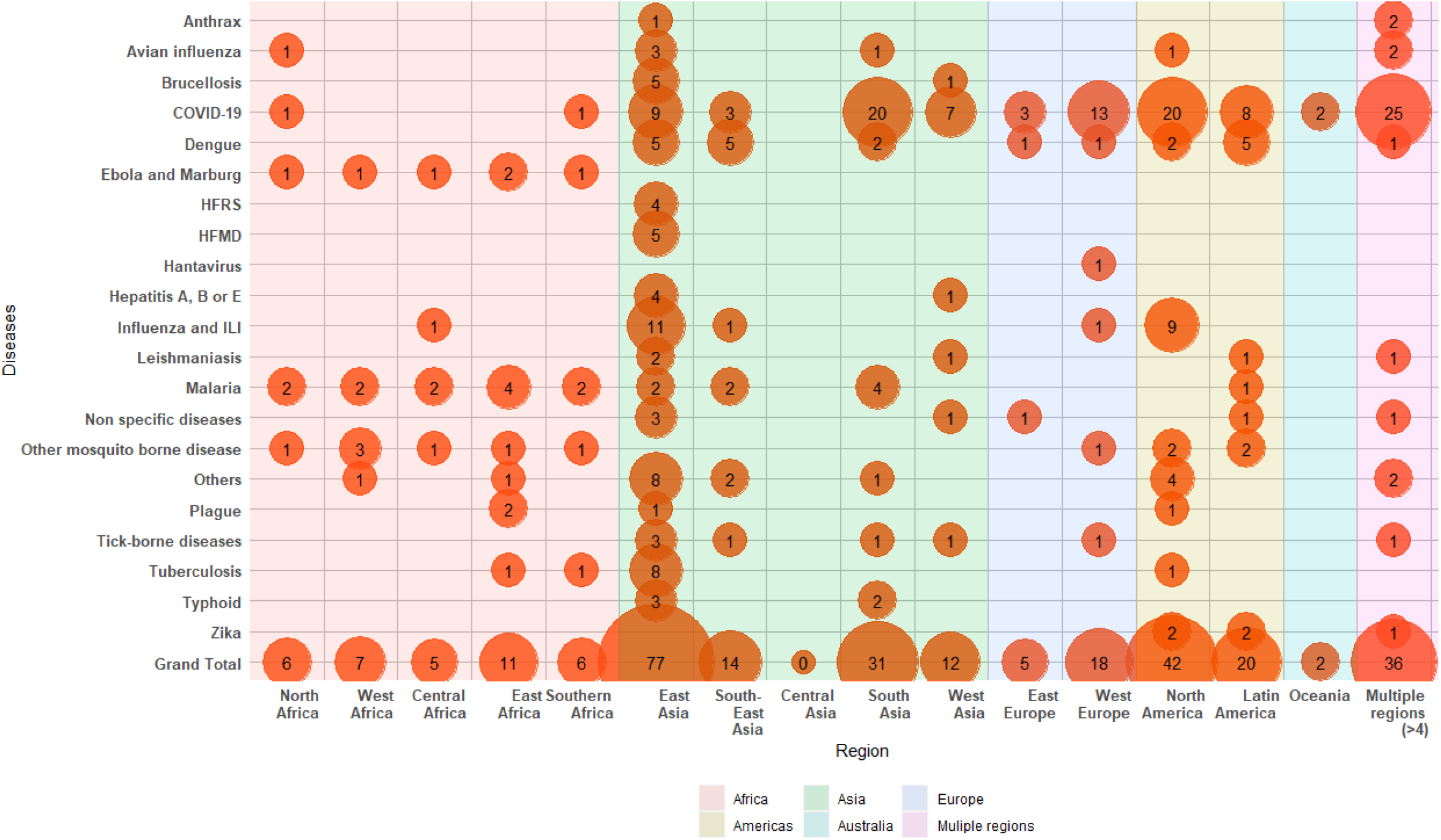
Distribution of articles with infectious disease models built for each geographical region. If an article included infectious disease models for more than four regions, they were placed in “multiple regions” category. Similarly, if an article included models for multiple diseases, they were placed in each respective category.

### Trend and extent of use of ML and DL in infectious disease prediction models

There has been an increasing trend in the use of ML and DL techniques for ID prediction since 2001 with a substantial rise between January 2019 and May 2021 (Fig. 4). Of the 237 articles included in the study, 127 (53.6%) of them applied at least one type of ML approach and 129 (54.4%) used at least one DL technique for disease prediction (Fig 4a). For the DL models, the FNN (63, 26.6%), RNN (48, 20.3%), and DL hybrids/ensembles (27, 11.4%) were the most common approaches (Fig 4b). Tree-based methods (91, 38.4%) followed by SVM (36, 15.2%) and then likelihood-based methods (22, 9.3%) were the most common ML approaches (Fig 4c). Within tree-based ML methods, Random Forest (RF) (44, 18.6%) followed by Boosted Regression Trees (BTR) (30, 12.7%) and Extreme Gradient Boosts (XGB) (12, 5.1%) were most often used (Fig 4d). More details including the citations groups by model type and subtype and are presented in Supplementary Table S1.

**Figure 4:**
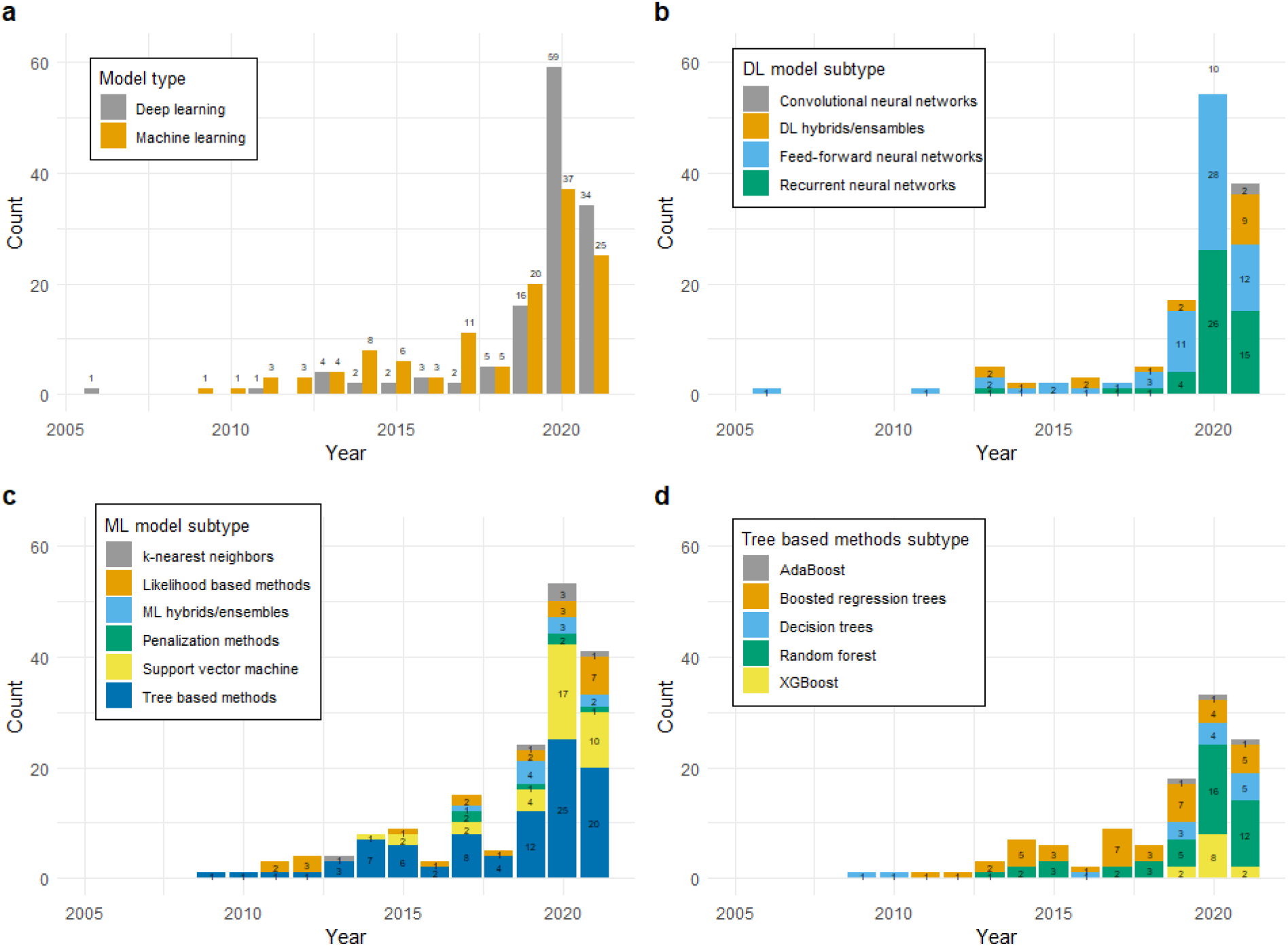
Trend and extent of ID prediction models published (January 2001-May 2021): Number of citations placed by **a)** model types (i.e., ML or DL) **b)** DL model subtypes **c)** ML model subtypes **d)** Tree-based ML model subtypes. Note: if an article contained models from different types or subtypes, it was placed in each respective group.

### Utilization of ML and DL approaches for different prediction categories

We grouped the 237 articles into prediction categories based on the tasks they performed. Majority of the articles performed temporal predictions (158, 66.7%) followed by disease risk predictions (90, 38.0%) and spatial predictions (74, 31.2%). COVID-19 was the most frequently modeled disease with the majority being temporal prediction models (Fig. 5). More details, including the citations groups by prediction categories and model type, are presented in Supplementary Table S1.

**Figure 5:**
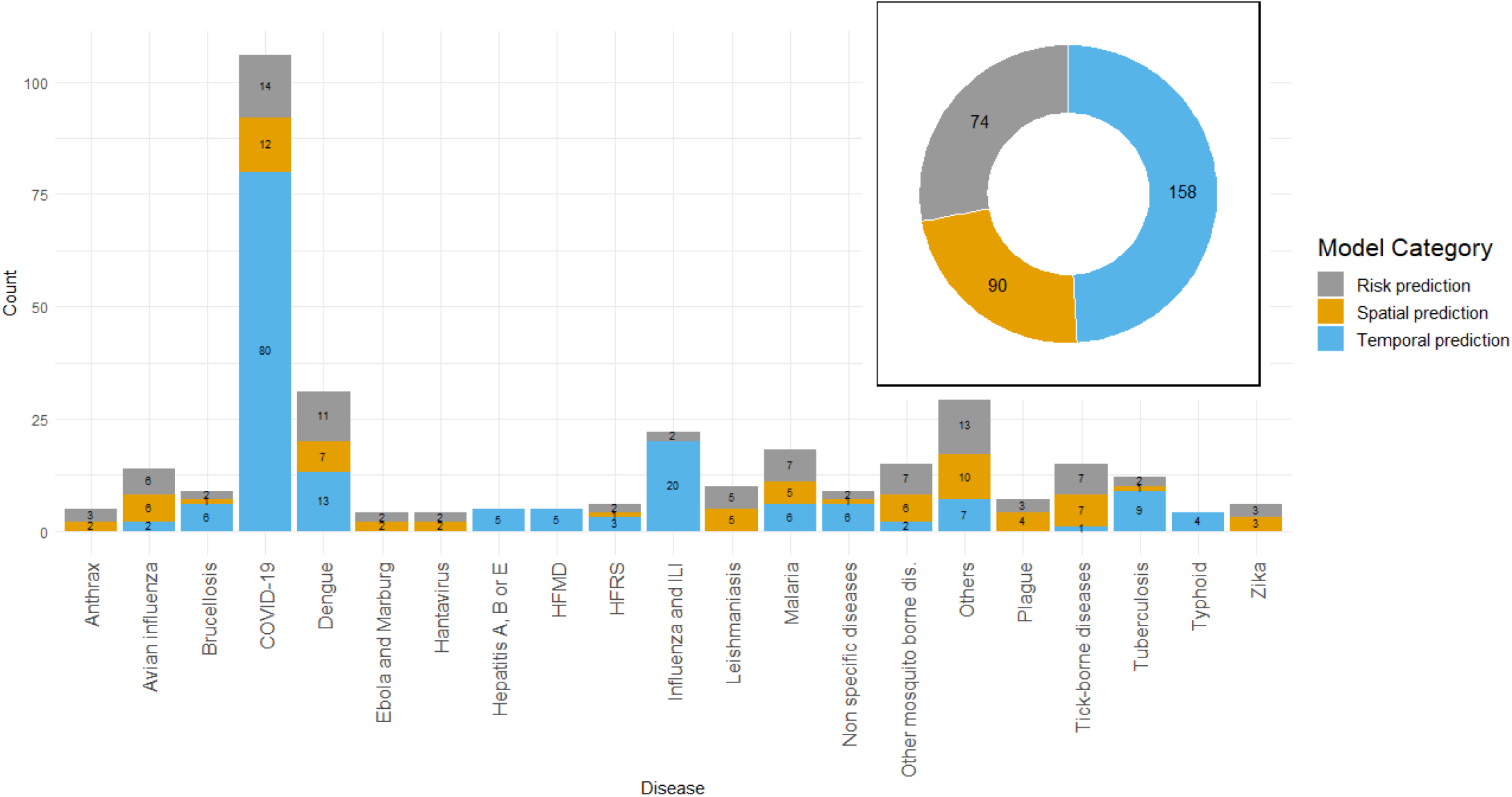
Model prediction categories. The distribution of disease prediction models grouped by model categories and diseases. If an article contained models that performed multiple prediction tasks and for multiple diseases, it was placed in each respective group.

### Spatial and temporal scales of the dataset used in the studies

The spatial scale (geographic extent) and temporal scales (duration) of the datasets used to make predictions were identified through the geographic extent/size and duration of the studies, respectively. Overall, for all ID prediction model categories, most articles predicted ID at the country level (124, 52.3%) using only up to one year of data (132, 55.7%) (Fig. 6a, 6b). Among temporal prediction models, near-term forecasting (up to one month) using one year worth of data was the most common (53, 33.5%) (Fig. 6c).

**Figure 6:**
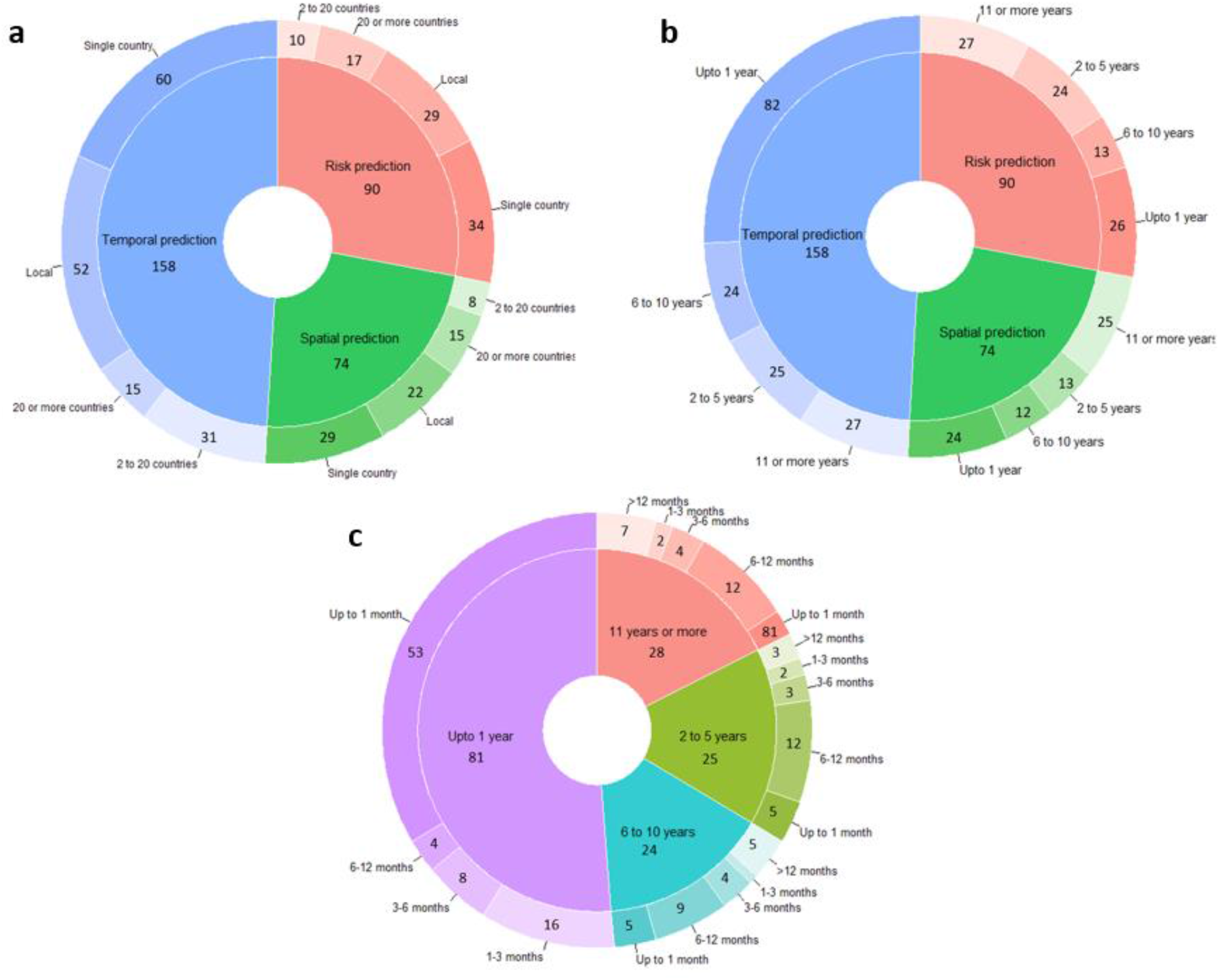
Spatial and temporal scale of ID prediction models. **a)** Proportion of the spatial scale (geographic extent) of the models grouped by model categories **b)** Proportion of temporal scale (duration) of the models grouped by model categories **c)** Among temporal prediction models, proportion of forecasting distance grouped by temporal scale. An article was placed in its respective groups if it utilized ID models with multiple model categories, spatial and/or temporal scales.

### Input feature groups utilized for disease prediction

The articles included in the study utilized input features that belonged to the following eight groups: case counts (154, 65.0%), climate/weather (98, 41.4%), demographics/socioeconomics (63, 26.6%), landscape/geography (58, 24.5%), social media/internet searches (18, 7.6%), health and comorbidity (7, 3.0%), human mobility (4, 1.7%), and news (3, 1.3%). Each disease modeled has a unique signature of input feature groups used for prediction (fig 7a). Focusing on the model prediction type categories, the number of input feature groups used in each category ranged from a minimum of one feature group (n = 151, 63.7%) to a maximum of five groups (n = 3, 1.3%) (fig 7b). A complete breakdown of the characteristics of each input feature group utilized for ID prediction is presented in Figure 3.

**Figure 7:**
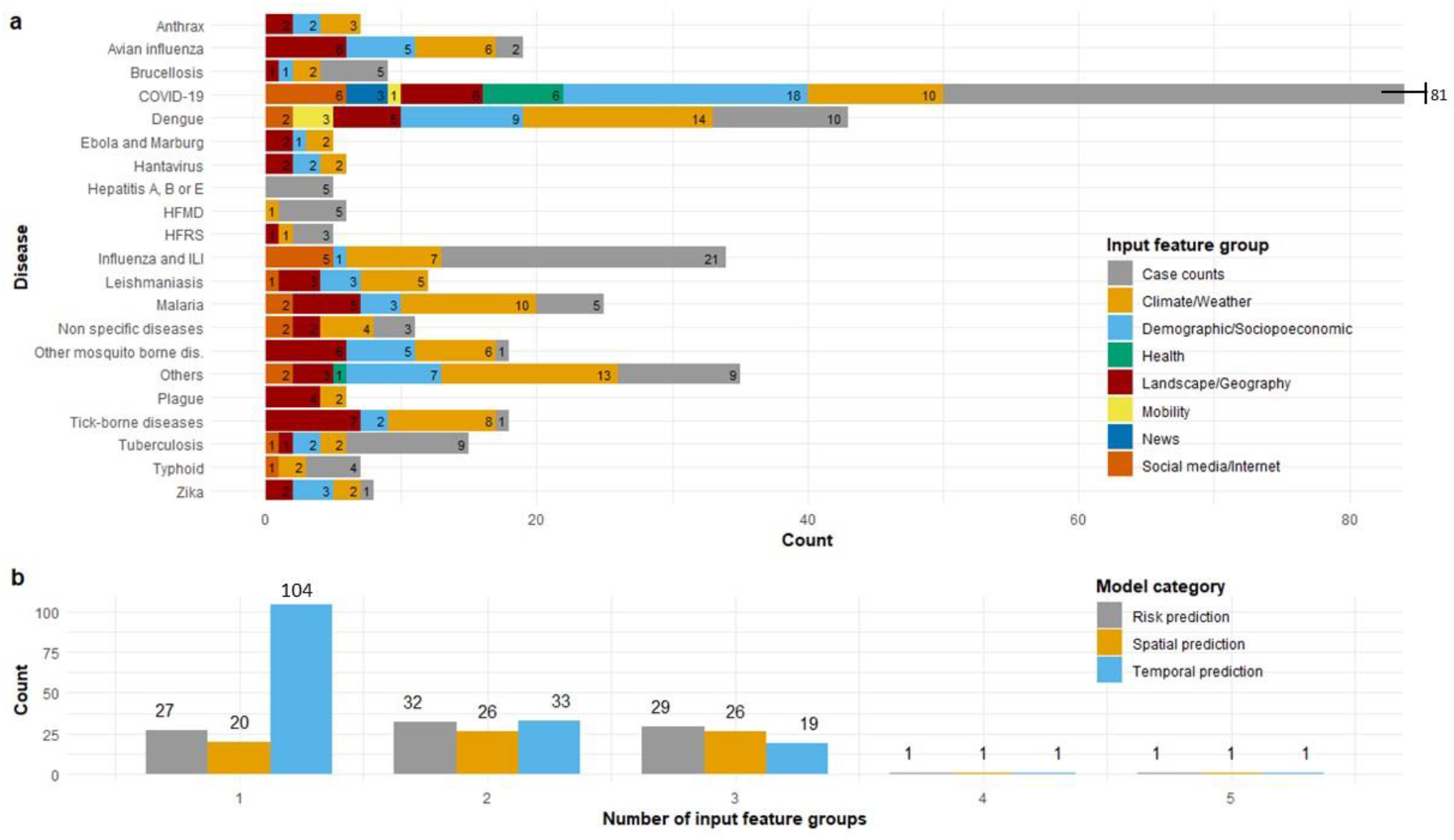
Characteristics of input feature groups utilized for disease prediction. Articles (n = 237) categorized by **a)** input feature groups used by disease type **b)** number of input feature groups utilized by ID prediction model categories. If an article utilized multiple input features, modeled multiple diseases and/or belonged to multiple model categories, the article was counted within each respective grouping.

### Uncertainty quantification, computational efficiency, and missing data

We identified only 21 (8.9%) of the articles to quantify uncertainty in their model predictions. The uncertainty quantification techniques used included frequentist (10, 4.2%)^a46, a67, a68, a91, a107, a123, a145, a152, a193, a195^, simulation/sampling based (7, 3.0%)^a26, a53, a156, a200, a213, a214, a219^, and Bayesian techniques (3, 1.3%) ^a94, a111, a115^.

Only 7 (3%) publications^a10, a13, a22, a63, a64, a79, a102, a103^ meeting the review criteria included information about computational efficiency while evaluating the performance of their models.

We also noted any missing data handling techniques used in model building. The majority of the articles (220, 84.4%) either did not report any missing data or did not explicitly mention how missing data was handled in their work. For the 18 (7.6%) articles that did discuss this topic, the techniques applied included replacement with mean/median or zeros^a56, a64, a72, a185, a187^, moving average^a136, a96, a128^, regression^a103, a108, a185^, correlation^a220^, KNN^a103^, multivariate imputation^a111, a136, a139^, exclusion^a24^, and pixel gap filling ^a157^.

### Common error metrics used in ID prediction modeling

Among classification models that predicted discreet values (e.g., presence or absence of a disease), Area Under the Curve - Receiver Operating Characteristic (AUC-ROC) curve (46, 19.4%), accuracy (29, 12.2%), and sensitivity (16, 6.8%) were the top three error metrics (Fig. 8b). Alternatively, among regression models that predicted continuous values (eg., monthly number of disease cases), Root Mean Square Error (RMSE) (98, 41.4%) followed by Mean Absolute Error (MAE) (67, 28.3%) and Mean Absolute Percentage Error (MAPE) (57, 24.1%) were the most common (Fig. 8b).

**Figure 8:**
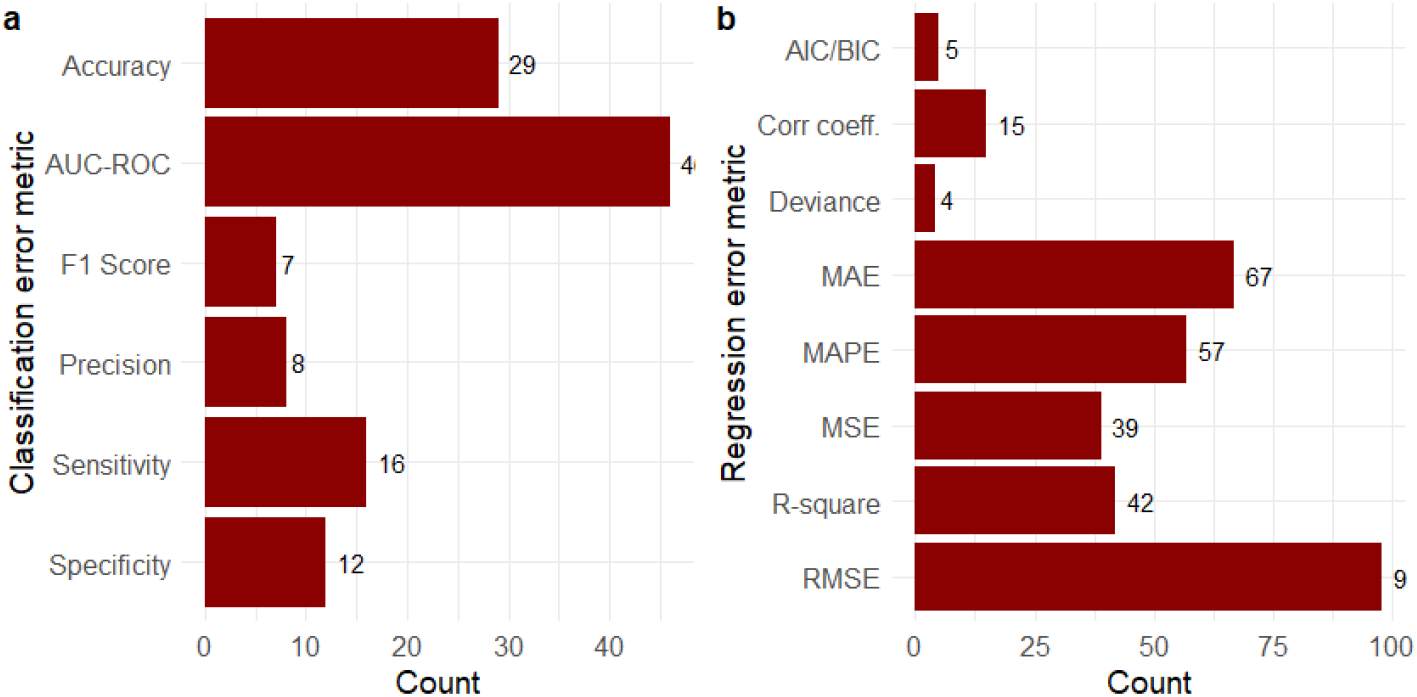
Error metrics utilized in ID prediction models: Citations grouped by **a)** Classification error metrics and **b)** Regression error metrics. If an article used error metrics from different classes, it was placed in each respective group. Abbreviations: AUC-ROC (Area Under the Curve - Receiver Operating Characteristic curve), AIC/BIC (Akaike’s/Bayesiasn Information Criteria, corr coeff. (Correlation coefficient), MAE (Mean Absolute Error), MAPE (Mean Absolute Percentage Error), MSE (Mean squared error), RMSE (Root Mean Square Error)

## Discussion

The ID threat is constantly changing across space and time, hence, an accurate and timely estimation of their occurrence is critical to planning and implementing successful disease preparedness and response strategies ^24,25^. This systematic review was conducted to understand the current state and extent of utilization of ML and DL algorithms in ID prediction. Our review showed that overall, there was a constant increase in the number of studies that utilized ML and DL to build ID prediction models between 2005 and 2019. Unsurprisingly, we saw an exponential rise in this trend after the COVID-19 pandemic outbreak. The overall global responses to the COVID-19 pandemic by the scientific community, governments, and non-government agencies have been unprecedented. This collective effort has resulted in increased collaboration among health sectors, large-scale disease surveillance, accessible data, and artificial intelligence technology sharing initiatives^26^. The availability of the crucial epidemiological knowledge through these initiatives along with the need for an accurate assessment of the disease dynamics has led to a dramatic increase in the utilization of ML and DL prediction modeling.

Most of the IDs that were modeled were either zoonotic in nature or diseases solely affecting the human population. Apart from COVID-19, influenza and influenza-like illnesses, dengue, malaria, and tuberculosis received major attention. These diseases have an ability to spread easily among the human community either directly through aerosolization or contact (influenza and influenza-like illnesses, tuberculosis) or propagated by vectors (dengue, malaria). This potential to spread easily and the ability to cause wide-scale mortalities and morbidities most likely led to increased attention from global health communities. Furthermore, almost all recent pandemics and a large proportion of emerging IDs originated from wildlife spillover and involve complex dynamic interactions within human and domesticated animal populations^27^. Our review found the majority of zoonotic diseases modeled had humans as their primary host. More efforts are required to integrate other host species that might significantly affect the transmission and persistence of an IDs across time and space. We identified only a very small number of publications on non-zoonotic livestock diseases, which could be due to inadequate livestock disease surveillance and the unavailability of reliable epidemiological data for modeling purposes. More efforts should be made to better predict these economic significance veterinary diseases since many of them, such as African swine fever, are highly contagious transboundary diseases with significant global food security and safety impacts.

The articles identified were almost evenly split between ML and DL techniques for ID prediction tasks. Within ML techniques, tree-based methods were popular among all prediction categories. Tree-based methods such as RF, BTR, and XGB are often among the best performing types of prediction models^9,28,29^. These models are also easy to implement, fast to compute, highly performant, and provide a form of interpretability through input feature importance, which could be the main reasons for their popularity in ID modeling^30,31^. Alternatively, FFNs, and RNNs were the most frequently used DL techniques and were mostly used for temporal prediction. The FFNs are artificial neural networks that can learn complex and non-linear patterns without making any prior assumptions concerning data distributions^32,33^. The RNNs are the derivatives of FFNs (e.g., Long Short-Term Memory and Gated Recurrent Unit) and are known to produce strong predictions with time series or other types of sequential data because of their ability to utilize historic information to predict future values^34^. Given that the ID outbreaks generally follow a non-linear and complex pattern, these neural networks are often shown to produce superior predictions compared to other approaches and are hence commonly used in disease forecasting tasks. It is also worthwhile to note that ML and DL hybrids/ensembles have attracted great attention from the ID communities in the past few years, evident by their increased use in publications. Hybrid and ensemble models are information fusion concepts that combine statistical, mechanistic, ML, and/or DL approaches working together (hybrid) or independently (ensemble) to minimize prediction noise and increase accuracy over the individual models, which could be one of the possible explanations for their increased popularity in recent years^8,35^.

A wide variety of input features were used for training ID models. Conventional variables (e.g., previous case counts, climate/weather, demographics/socioeconomics, and landscape/geographic data) were routinely utilized to make disease predictions. However, one of the biggest constraints for building a reliable ID prediction model to accurately estimate the progression of the disease is the timeliness of available, essential outbreak-related data. These constraints are aggravated in cases involving a novel disease outbreak or neglected endemic disease where the spatial and temporal patterns of the pathogen emergence are largely unknown. Furthermore, a major outbreak could lead to a significant shift in population social behavior and movement due to public health efforts and government policies resulting in prediction inaccuracies. Hence, the incorporation of novel data sources that account for these dynamic behaviors is vital for accurate and timely decision making. In our review, we identified studies that utilize news articles, social media or internet search queries, heath information collected using phone/wearable devises, and human mobility data. The ML and DL models used in these studies exploited a large quantity of structured and unstructured data with the goal to produce better ID predictions.

Although methods used in ID prediction are becoming more sophisticated, we also identified consistent concerns in the structure of the analyses that could limit their practical use. First, we found that the data collection duration for a large majority of the studies was less than or equal to one year regardless of the prediction category. Secondly, most articles do not include uncertainty quantification or account for missing data. This was apparent especially during the early stages of the COVID-19 pandemic where the availability of data was limited and there was a widespread underreporting of the cases. Since each ID is known to show specific occurrence patterns that change over time and space, such short-term predictions could be subject to biases and estimation inaccuracies, which should be carefully accounted for while deploying the algorithms to an operational environment. Though a short turnaround time could be vital for a real-world ID event, we recommend updating the models regularly with new data and retraining them for better and long-term practical usage.

Another serious limitation common to the literature reviewed is a lack of discussion regarding data quality and the functional deployment of an algorithm. While one algorithm may perform the best in terms of overall tested accuracy, it may overstate its confidence, may be unrealistic to implement due to computational efficiency, or may simply fail when in the presence of missing data. Since disease prediction models are meant to provide situational awareness, reliable and near-real-time results are necessary^36^. The fact that so few publications consider the critical aspects of automated algorithm implementation suggests that a greater emphasis should be placed on the operational aspects of epidemiology for operational biopreparedness and response.

While our systematic review was comprehensive, it still has some limitations. First, we only included peer-reviewed studies that were published in a scientific journal. This could have resulted in a selection bias by excluding important studies disseminated as preprints, conference proceedings, books, dissertations, or theses. Second, we did not include studies that primarily utilized traditional statistics-based regression or classification methods. Considering the amount of literature available about these techniques, they will require a separate literature review of their own.

In summary, we conducted a systematic review to determine the current state of ID prediction capabilities that utilized ML and DL techniques. We specifically looked for IDs that were modeled, type of the ML and DL techniques utilized, the geographical distribution of the modeling studies, prediction tasks performed, input features utilized, spatial and temporal scale of the studies, error metrics used, the computational efficiency of the models, uncertainty quantification and missing data handling methods adapted. We observed a diverse number of IDs modeled with COVID-19 appearing most frequently in recent literature. We also note that there has been a consistent increase in the number of studies that apply use ML and DL techniques in ID prediction tasks over the past two decades. However, despite the increased use of data-driven methodology, more studies are needed that include the full disease ecology specifically for zoonotic and veterinary disease predictions from the human, animal, vector, and environmental aspects in a One-Health context. Finally, to enable biopreparedness and response, studies should incorporate the assessment of uncertainty in their predictions and computational requirements of their models, which are crucial for operational deployment.

## Data Availability

All data generated or analyzed during this review are included in this article and its supplementary information files

## Acknowledgements

The authors wish to thank Nakita Pradhan, Samuel Ortega, and Jaidyn Bryant for their contribution in the initial review process. The authors wish to thank Samantha Erwin for reviewing the manuscript and providing general feedback. RK acknowledges the support from the Pacific Northwest National Laboratory (PNNL)-Washington State University (WSU) Distinguished Graduate Research Program (DGRP) Fellowship for facilitating this research collaboration.

## Author contributions

R.K., L.E.C., K.P. and S.D. designed the search strategy, implemented the study protocol, and retrieved articles. R.K. led the screening and data extraction process, performed data analysis and visualization of the results. R.K., L.E.C., K.P., wrote the manuscript. L.E.C and K.P. acquired the funding. All authors reviewed the manuscript and agreed to the published version of the manuscript.

