## Supplementary Table S1 for "Predicting infectious disease for biopreparedness and response: A systematic review of machine learning and deep learning approaches"

**SUPPLEMENTARY INFORMATION**

Predicting infectious disease with artificial intelligence: A systematic review of machine learning and deep learning approaches

Ravikiran Keshavamurthy[1,2,] Samuel Dixon[1], Karl T. Pazdernik[1,3] and Lauren E. Charles[1,2 *]

[1]Pacific Northwest National Laboratory, Richland, WA 99354, USA

[2]Paul G. Allen School for Global Health, Washington State University, Pullman, WA 99164, USA

[3]Department of Statistics, North Carolina State University, Raleigh, NC 27695, USA

**Supplementary Note**

Supplementary Table S1: Citations categorized by prediction category, model type, and subtype with corresponding number of articles for each.

| **Model prediction categories** | **Model type (no. articles, % category** | **Model subtype** | **No. of articles** |
|---|---|---|---|
| Temporal prediction | ML | Tree-based models [a4, a7, a23, a45, a54, a57, a58, a70, a72, a75, a80, a90, a92, a100, a104, a106, a107, a112, a116, a122, a126, a138, a157, 1a63, a167, a180, a181, a183, a185, a186, a188] | 31 |
| | | Support Vector Machines [a4, a6, a8, a23, a40, a44, a54, a57, a70, a73, a75, a80, a90, a106-108, a120, a122, a124, a141, a176, a181, a183, a185, a186, a190, a197] | 27 |
| | | ML Hybrid/Ensemble[a17, a34, a64, a81, a99, a119, a190, a204] | 8 |
| | | Penalization methods[a4, a54, a91, a94, a113, a124] | 6 |
| | | Likelihood-based[a2, a12, a50, a73, a193] | 5 |
| | | k-nearest neighbor[a70, a80, a182] | 3 |
| | DL | FNN[a1, a10, a13, a14, a19, a21, a25, a30, a31, a33, a35, a41, a45, a48, a50, a52, a57, a62, a66, a71, a73, a75, a87, a94, a95, a97, a102, a115, a120-122, a124, a126, a127, a135-137, a149, a168, a177, a178, a181, a183, a186, a194, a205, a210] | 48 |
| | | RNN[a5, a6, a15, a16, a18, a28, a29, a30, a32, a39, a40, a42, a43, a45, a46, a51-53, a55, a60, a61, a66, a67, a78, a79, a85, a86, a88, a90, a93, a96, a98, a101-103, a105, a122, a125, a137-139, a150, a153, a186, a191, a192, a196, a197, a210] | 48 |
| | | DL Hybrid/Ensemble a3, a16, a22, a26, a37, a38, a47, a49, a64, a68, a69, a82, a84, a110, a118, a144, a145, a146, a151, a152, a179, a187, a189, a195, a225, a227, a231 | 27 |
| | | CNN[a16, a45, a53, a60, a101, a186, a191] | 7 |

| | | | |
|---|---|---|---|
| Spatial prediction | ML | Tree-based models[a7, a20, a36, a57, a83, a111, a114, a130, a132, a140, a142, a143, a156-162, a164-166, a169-175, a184, a199-201, a207-209, a211, a213, a214, a216, a217, a218, a219, a221, a222, a224, a228, a232-234, a236, a237] | 52 |
| | | Likelihood[a131, a133, a142, a155, a171, a199, a203, a206, a209, a215, a216, a232, a236] | 13 |
| | | Support Vector Machines[a36, a57, a124, a140, a142, a199, a209, a232, a236] | 9 |
| | | ML Hybrid/Ensemble[a34, a64, a199] | 3 |
| | | k-nearest neighbor[a36, a228] | 2 |
| | | Penalization methods[a124] | 1 |
| | | FNN[a27, a36, a57, a124, a140, a148, a158, a177, a198, a219, a220, a226, a236] | 13 |
| | | RNN[a79, a125] | 2 |
| | | DL Hybrid/Ensemble[a22, a64] | 2 |
| Disease risk | ML | Tree-based models[a20, a36, a45, a58, a59, a65, a83, a109, a111, a114, a116, a123, a126, a128-130, a132, a134, a140, a142, a143, a147, a156-162, a164-166, a169-175, a181, a183, a184, a199-201, a207-209, a211, a213, a214, a216-219, a221, a222, a224, a228, a229, a230, a232-237] | 67 |
| | | Likelihood[a89, a128, a129, a131, a133, a142, a155, a171, a199, a203, a209, a212, a215, a216, a232, a236] | 16 |
| | | Support Vector Machines[a36, a128, a129, a140, a141, a142, a181, a183, a199, a209, a232, a236] | 12 |
| | | ML Hybrid/Ensemble[a17, a56, a199] | 3 |
| | | k-nearest neighbor[a36, a228, a229] | 3 |
| | DL | FNN[a11, a27, a36, a45, a126, a128, a140, a141, a148, a154, a158, a177, a181, a183, a198, a219, a220, a223, a235, a236] | 20 |
| | | RNN[a18, a45, a67] | 3 |
| | | CNN[a45] | 1 |
| | | DL Hybrid/Ensemble[a38] | 1 |
